# WHO SHOULD BEAR THE COST OF COVID-19 NON-VACCINATION? A LUCK EGALITARIAN ASSESSMENT IN A SOUTH AFRICAN INSURED POPULATION

**DOI:** 10.64898/2026.07.07.26357455

**Authors:** Geetesh Solanki, Francesca Little, Susan Cleary

**Affiliations:** Health Economics Unit, Faculty of Health Sciences, Anzio Road, Observatory, 7925, Cape Town, South Africa; Department of Statistical Sciences, University of Cape Town, South Africa

**Keywords:** Luck egalitarianism, Personal responsibility, COVID-19 vaccination, Health insurance, Cross-subsidisation, Fairness and solidarity, South Africa

## Abstract

**Background:** Personal choice in health behaviours raises difficult questions: when individuals freely decline effective preventive interventions, who should bear the resulting costs? This tension is acute in insurance systems where resources are pooled, yet all health systems pursuing Universal Health Coverage must navigate the boundary between collective solidarity and individual accountability. During the COVID-19 pandemic, vaccines were freely available to members of South African private medical schemes, creating conditions in which non-vaccination could plausibly be examined as a matter of personal choice rather than constrained access. This study applied a luck egalitarian framework to assess whether non-vaccination reflected personal choice or constrained circumstance, and to quantify resulting excess costs.

**Methods:** A contextual review assessed barriers to vaccination. Using de-identified claims data for approximately 550,000 individuals (March 2020 to December 2022), logistic regression estimated each person’s predicted probability of vaccination based on demographic and clinical factors, with observed and predicted rates compared across strata to infer choice versus circumstance. A zero-inflated negative binomial model estimated predicted expenditure among vaccinated members, applied to the full population to simulate universal vaccination. Excess costs were calculated across predicted probability strata.

**Results:** Predicted and observed vaccination rates were closely aligned, suggesting that residual non-vaccination in higher-probability groups reflected personal choice rather than constrained circumstance. Observed costs exceeded predicted costs by 22% under universal vaccination, concentrated among older adults and those with comorbidities. Among those with a 60 to 70% predicted probability of vaccination, observed costs exceeded predicted costs by 127.6%. In contrast, among younger, low-risk members, predicted costs slightly exceeded observed expenditure, as vaccination costs were not offset by reduced hospitalisation.

**Conclusion:** Risk pooling depends on solidarity, yet non-vaccination due to personal choice shifts costs in ways that challenge fairness in community-rated insurance. These findings highlight the need for transparent deliberation about when personal responsibility should inform equitable health financing design.

**KEY MESSAGES:**

- COVID-19 vaccination was freely accessible to the South African insured population, with minimal financial or structural barriers.
- Predictive modelling helped distinguish individuals whose non-vaccination was more likely driven by choice rather than circumstantial barriers.
- Unvaccinated members generated substantially higher COVID-19–related healthcare costs than expected under a universal vaccination scenario, particularly among older adults and those with comorbidities.
- These excess costs were collectively absorbed by the risk pool, resulting in a cross-subsidy from vaccinated to unvaccinated members.
- Transparent deliberation is needed on when personal responsibility should inform health financing design within solidarity-based insurance arrangements.

## Introduction

People make different choices about how to live their lives. Personal choice is understood here as an individual’s opportunity and autonomy to perform an action selected from at least two available options, unconstrained by external parties(Cappelen et al., 2005). It encompasses health-related behaviours or decisions that are, or could reasonably be expected to be, within an individual’s control.

Health-related choices affect individuals’ health, the risks they face, and their need for treatment (Cappelen et al., 2006). A large body of evidence shows that unhealthy lifestyles contribute significantly to the high and growing burden of disease globally (2017, Vos et al., 2020, Ferretti, 2015). As a result, scarce medical resources that could be directed at preventing or treating suffering for which no individual is responsible are instead consumed by conditions that could be avoided through individual behaviour (Farrenkopf, 2022).

Personal choice-driven increases in disease burden are placing growing demands on resource-constrained health care systems globally (2017, Vos et al., 2020, Ferretti, 2015). This has encouraged theorists and decision makers to ask whether individuals should be required to take greater personal responsibility for their health. In this context, personal responsibility refers to any health policy that prioritises services by linking either the relative payment for treatment or the extent of treatment to factors that are under an individual’s control. The underlying logic is that those who make personal choices that increase their risk of illness or injury should bear a greater share of the associated costs or face limitations on the care they receive (Feiring, 2008, Cappelen et al., 2005).

The question of personal responsibility is particularly relevant in South Africa, where behavioural risk factors contribute substantially to the country’s disease burden (Achoki et al., 2022, Matzopoulos et al., 2022, Bradshaw et al., 2022, Neethling et al., 2022, Groenewald et al., 2022). While South Africa has committed to Universal Health Coverage (UHC), underpinned by principles of access, equity, quality, and financial risk protection, these values may be compromised by the pragmatic measures required to deliver even a modest version of UHC. As the country moves towards National Health Insurance (NHI), it will be critical to establish a clear set of benefits and a framework for selecting them. If quality care cannot be provided for all needs, what needs should be prioritised, and on what basis? At what point is it legitimate to shift responsibility from the collective towards individual citizens?

There is a case, therefore, for considering whether personal responsibility should be included as a prioritisation criterion in benefit package design. However, applying personal responsibility in health is complex and highly contested (Sharkey et al., 2010), requiring careful assessment across multiple dimensions, including impact, fairness, and practicality. While the need for such assessment is clear, the literature offers limited practical guidance on how it should be conducted, and empirical case studies remain rare.

A philosophical framework that has gained prominence in these debates is luck egalitarianism (Cappelen et al., 2005, Cappelen et al., 2006). Luck egalitarianism distinguishes between inequalities arising from factors beyond an individual’s control, referred to as “brute luck” and those arising from genuine personal choices, referred to as “option luck”(Cappelen et al., 2005, Dworkin, 1981b, Segall, 2010). According to this view, society has a duty to compensate individuals for disadvantages resulting from brute luck, but inequalities resulting from option luck may be considered acceptable and individuals should bear the consequences of their genuine choices (Albertsen et al., 2014, Voigt). Roemer operationalised this distinction by separating outcomes due to “circumstances” (factors beyond control) from those due to “effort” (matters of personal responsibility).

COVID-19 vaccination in South Africa’s privately insured population offers an interesting case study for applying this framework. During the pandemic, vaccines were provided free of charge, administration costs were covered by medical schemes, and vaccine procurement was funded by the state (CMS, 2021a, CMS, 2022). These features removed financial risk protection barriers, and the widespread availability of vaccination sites addressed physical access. Extensive communication campaigns by schemes and public authorities further addressed information-related acceptability barriers, though unmeasured dimensions such as vaccine hesitancy and trust may have persisted. Together, these conditions created a setting in which non-vaccination could more plausibly be attributed to personal choice rather than to financial, physical, or informational constraints on access. The insured population thus serves as a useful “pseudo-laboratory” for operationalising luck egalitarian principles in a real-world context. This setting allows us to ask: when individuals chose not to vaccinate despite free and widespread availability, should they bear the resulting health care costs?

This study addresses the gap between theoretical debates on personal responsibility and empirical evidence. Using COVID-19 vaccination in a South African insured population as a case study, it asks: when individuals choose not to vaccinate despite free and widespread availability, should they bear the resulting health care costs? We pursue two aims: first, to assess whether non-vaccination reflected genuine personal choice rather than constrained circumstance; and second, to quantify the distribution of COVID-19-related costs between vaccinated and unvaccinated members of the insurance pool. In doing so, the study provides an empirical application of luck egalitarian principles to a real-world health financing question.

## METHODS

### Overview

This study draws on secondary insurance claims data to operationalise a luck egalitarian framework in the context of COVID-19 vaccination within a South African insured population. The analytical approach is informed by the work of John Roemer, who developed a formal framework for distinguishing outcomes due to “circumstances” (factors beyond individual control) from those due to “effort” (matters of personal responsibility). Roemer’s method involves dividing the population into “types” sharing the same circumstances and comparing effort within types. While the available data limited us to a regression-based approach rather than Roemer’s full “type” methodology, the underlying logic is the same: individuals with a high predicted probability of vaccination (given their circumstances) who nevertheless remain unvaccinated are inferred to have made a genuine personal choice.

Following the principles of the STROBE guidelines(Vandenbroucke et al., 2007) for observational studies and the CHEERS 2022 guidelines(Husereau et al., 2022) for economic evaluations, we sought to ensure transparent reporting of study design, data, and analysis, while recognising that the hybrid normative–empirical nature of this study does not map perfectly onto standard reporting checklists.

The study comprised two components, corresponding to the two research aims: The first assessed whether non-vaccination could reasonably be attributed to personal choice rather than constrained circumstance, where circumstance is understood broadly to include factors beyond individual control, such as access barriers. This step is foundational: only if non-vaccination reflects choice can the resulting costs be fairly attributed to individual behaviour. The second component quantified the extent to which non-vaccinated members generated excess costs borne by the insurance risk pool as a whole. An overview of the data, the analytic approach, the outputs and interpretation are summarized in Figure 1.

**Figure 1.**
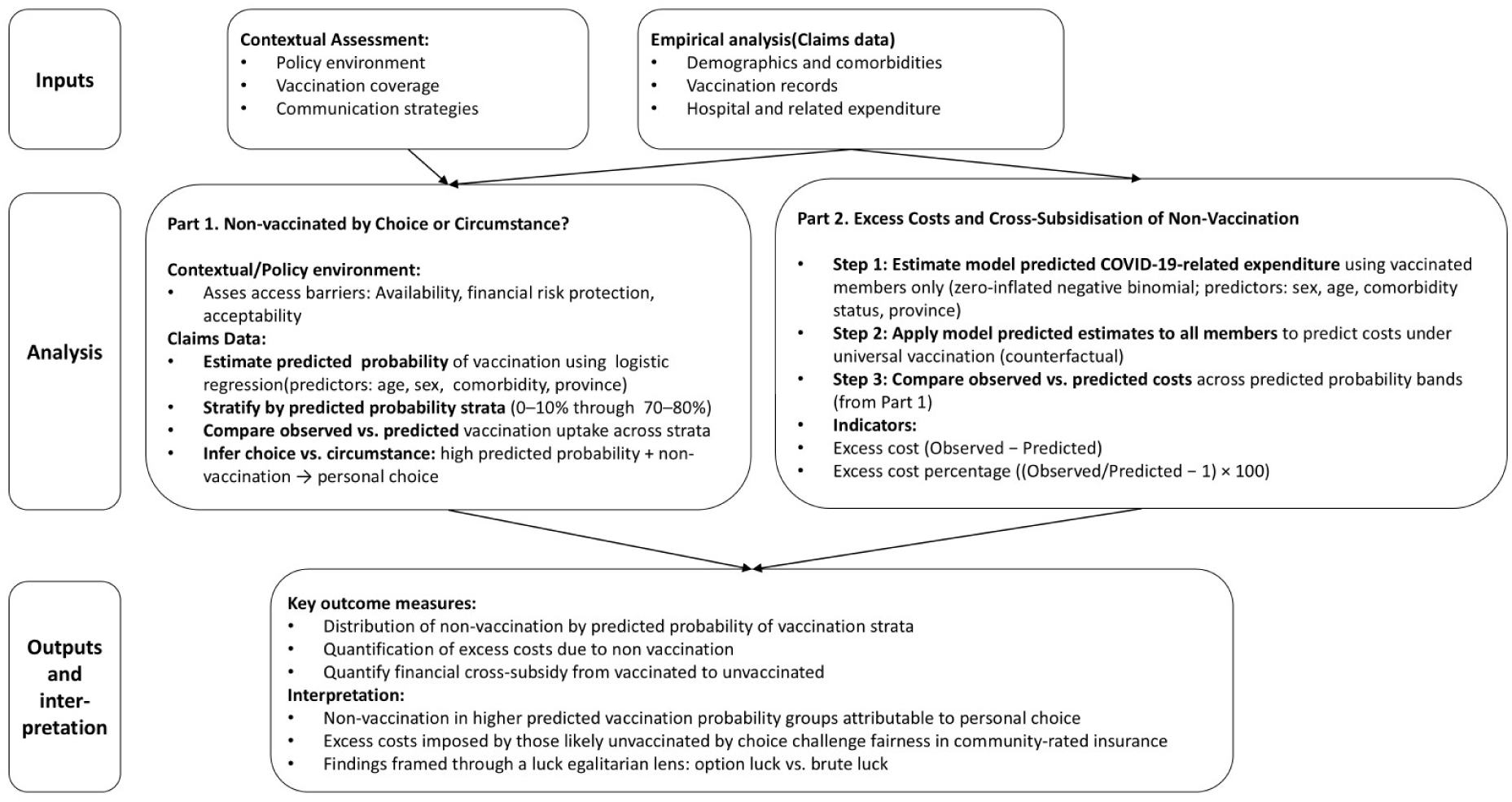
Overview of data inputs, analytic approach, and outputs and interpretation.

### Setting, Context, and Study Population

The study population consisted of 550,332 individuals who were enrolled in one of two large medical schemes between 1 March 2020 and 31 December 2022. This period corresponds to the first confirmed COVID-19 case in South Africa (5 March 2020) and the end of the fourth wave with the lifting of restrictions (30 December 2022). The sample represents approximately 5% of the South African insured population and is broadly comparable to the national insured population in terms of household structure, contributions, and health expenditure patterns (Solanki et al., 2025).

Vaccination coverage was determined by national rollout policy. Two vaccines were primarily used in South Africa during the study period: the Johnson & Johnson (J&J) adenoviral vector vaccine and the Pfizer-BioNTech mRNA vaccine (Bekker et al., 2022, Collie et al., 2022). Vaccination status was defined dynamically over time, following definitions established in the literature (Havers et al., 2022) and consistent with South African practice (Discovery, 2021). For the analyses in this paper, individuals who received at least one dose of either vaccine were classified as vaccinated; those with no recorded doses were classified as non-vaccinated. While earlier research on this cohort distinguished between partial and full vaccination (Solanki et al., 2025), this paper’s analytical requirements, particularly the need to generate stable predicted probabilities and counterfactual cost estimates, favoured a binary classification. This also allowed for clearer interpretation of the choice-versus-circumstance distinction.

### Data Sources

De-identified individual-level claims data were extracted from the data warehouse of NMG Consultants and Actuaries, an independent actuarial firm managing the schemes included. Three linked datasets were constructed:

- Demographic dataset which consisted of a record per enrolled member, including unique study identifier, sex, age, province of residence, enrolment dates, and comorbidity indicators. Comorbidities considered were those identified as clinically relevant for COVID-19 outcomes (cancer, chronic renal disease, congestive cardiac failure, COPD, diabetes, HIV, hypercholesterolemia, hypertension, hypothyroidism, ischemic heart disease, pregnancy, tuberculosis). These were identified following Council for Medical Schemes (CMS) claims-based algorithms(Council for Medical Schemes).
- Vaccination dataset which consisted of a record for each COVID-19 vaccination event, including vaccine type, date, provider, and amount reimbursed. Vaccination status was defined dynamically over time and collapsed into an end-of-period measure: vaccinated (≥1 dose) vs. non-vaccinated.
- A hospital and related expenditure dataset which consisted of a record for each COVID-19 hospital admission, including admission/discharge dates, length of stay, ICD-10 diagnosis codes, procedure codes, and total reimbursed hospital expenditure. Individuals with multiple admissions contributed multiple records.

Total hospital and vaccination related expenditures took an insurer perspective and reflected the amounts paid by the health insurance funds to cover the claims submitted by health care providers for services rendered to the hospitalized individuals. These direct costs included health care providers, hospital bed days, as well as any pharmaceuticals, medical or surgical consumables dispensed, and vaccinations provided. All expenditures in South African Rands (ZAR) were converted to United States Dollar ($) values using the December $/ZAR exchange rate for the given year and then inflated/deflated to be expressed in 2022 values using the South African Consumer Price Index(StatsSA, 2024).

### Part 1: Assessing Non-vaccination as Choice versus Circumstance

We distinguished between non-vaccination due to personal choice (voluntary refusal despite available opportunity) and due to constrained circumstance where circumstance is understood broadly to include factors beyond individual control, such as access barriers.

For the contextual assessment, following established frameworks in the health care access literature (McIntyre et al., 2009), we conceptualised access barriers across three dimensions: availability, financial risk protection, and acceptability. The barriers across the three dimensions were assessed by a contextual review of the policy environment, vaccination coverage and communication strategies. We reviewed scheme-level benefit coverage, Council for Medical Scheme circulars mandating COVID-19 vaccination as a prescribed minimum benefit (CMS, 2022, CMS, 2021a), and publicly available scheme communication strategies during the rollout(Discovery, 2021).

Empirically, we estimated each member’s probability of vaccination using logistic regression with vaccination status (vaccinated vs. non-vaccinated) as the dependent variable. Independent variables included sex, age group (<18, 18–25, 25–40, 40–65, ≥65), presence of COVID-19 comorbidity (yes/no), and province of residence. The model produced a predicted probability of vaccination for each individual, reflecting the predicted uptake given their demographic and clinical profile.

To infer the relative role of choice versus circumstance, we then grouped individuals into strata based on their predicted probability of vaccination (0–10% to 70–80%), allowing comparison of actual and predicted vaccination rates across groups with similar predicted probability of vaccination. The underlying logic, informed by Roemer’s framework, is that individuals with a high predicted probability of vaccination (given their circumstances) who nevertheless remain unvaccinated can reasonably be inferred to have made a genuine personal choice. Conversely, those in low-probability strata may have faced unmeasured access barriers or other circumstances beyond their control. We did not set any arbitrary cut-offs, but persistent non-vaccination in higher-probability strata (e.g., ≥50%) is more plausibly attributable to personal choice, while non-vaccination in lower strata (<30%) could reflect “constrained circumstance” or “access barriers”.

### Part 2: Assessing Excess Costs and Cross-subsidisation

To quantify the excess costs associated with non-vaccination, we conducted a three-step analysis. In the first step, we restricted the dataset to vaccinated members and used their information to build a benchmark model to estimate the predicted COVID-19–related costs had the individual vaccinated. Because most individuals incurred no COVID-19 hospital expenditure while a smaller proportion incurred very high costs, we applied a zero-inflated negative binomial (ZINB) regression model. This model accommodates both the probability of zero costs and the over-dispersed distribution of positive costs. Predictor variables included sex, age group, comorbidity status, and province. The model generated a predicted cost for each member, representing the average expenditure anticipated for an individual with their risk profile if they had vaccinated.

In the second step, we applied this benchmark model to the entire study population, both vaccinated and unvaccinated, to generate a counterfactual scenario estimating what each individual’s costs would have been if they had been vaccinated, given their demographic and clinical characteristics. This produced predicted expenditure under a universal vaccination scenario.

In the third step, we compared observed and predicted costs across the predicted probability bands generated in Part 1, allowing direct comparison between actual expenditure and the counterfactual for each individual.

Two indicators were derived to quantify the additional costs associated with remaining unvaccinated. The excess cost was calculated as the absolute difference between observed and modelled (predicted) expenditure, while the excess cost percentage was calculated as ((Observed/Predicted)-1)*100, representing the proportional deviation from predicted expenditure. Positive values indicate higher costs than would be predicted under the universal vaccination scenario, denoting greater costs attributable to non-vaccination. Together, these indicators quantify both the absolute and relative extent to which higher costs among unvaccinated members increased the collective costs borne by the insured risk pool. Finally, we examined observed versus predicted expenditure and excess cost across strata defined by predicted vaccination probability bands (from Part 1 of the analysis).

## RESULTS

### Study Population

The study included 550,327 individuals enrolled between March 2020 and December 2022. Of all the individuals, 53.6% were female, 34.4% were less than 18 years old, 23.3% had one or more comorbidity, and the highest proportion (45.4%) of individuals resided in Gauteng province (Table 1).

**Table 1.** Characteristics of study population.

| Variable | Number of individuals |  |
| --- | --- | --- |
|  | N | % of total |
| **Sex** |  |  |
| Female | 295,112 | 53.6% |
| Male | 255,216 | 46.4% |
| **Age** |  |  |
| Less than 18 | 189,402 | 34.4% |
| Between 18-25 | 40,190 | 7.3% |
| Between 25-40 | 145,003 | 26.3% |
| Between 40-65 | 140,255 | 25.5% |
| Greater than 65 | 35,477 | 6.4% |
| **COVID-19 Comorbidities** |  |  |
| No | 422,116 | 76.7% |
| Yes | 128,215 | 23.3% |
| **Province** |  |  |
| Eastern Cape | 40,877 | 7.4% |
| Free State | 22,347 | 4.1% |
| Gauteng | 249,646 | 45.4% |
| KwaZulu-Natal | 85,572 | 15.5% |
| Limpopo | 19,410 | 3.5% |
| Mpumalanga | 18,514 | 3.4% |
| North-West | 19,064 | 3.5% |
| Northern Cape | 10,002 | 1.8% |
| Western Cape | 84,895 | 15.5% |
| **Total** | 550,327 | 100.0% |

### Part 1: Non-vaccination by Choice versus Circumstance

A contextual review of policy environment, vaccination coverage and communication strategies was undertaken to assess access barriers in terms of availability, financial risk protection and acceptability. In terms of financial risk protection, vaccines were provided free of charge to all eligible individuals, with costs of administration covered by medical schemes and vaccine procurement covered by the state (CMS, 2021a, CMS, 2021b). Access was supported through a network of public and private vaccination sites, and major schemes, including those analysed here, implemented extensive member communication campaigns via digital portals, SMS prompts, and call-centre support (Discovery, 2021). These features substantially reduced barriers linked to financial risk protection (no out-of-pocket costs) and availability (widespread vaccination sites). Extensive communication campaigns by schemes and public authorities further addressed information-related barriers, though unmeasured dimensions of acceptability, such as vaccine hesitancy and trust, may have persisted.

In terms of the empirical analysis, an overall 37.2% of the study population received at least one dose of a COVID-19 vaccine by the end of study period (Table 2).

**Table 2.** Univariate and multivariate analysis of vaccination probability.

| Variable | Number of individuals |  |  |  |
| --- | --- | --- | --- | --- |
|  | N | Number Vaccinated | % Vaccinated | Adjusted OR (95 CI) |
| **Sex** |  |  |  |  |
| Female | 295,112 | 116,328 | 39.4% (39.2%-39.6%) | Reference |
| Male | 255,216 | 88,288 | 34.6% (34.4%-34.8%) | 0.89 (0.88-0.90) |
| **Age** |  |  |  |  |
| Less than 18 | 189,402 | 13,547 | 7.2% (7.0%-7.3%) | Reference |
| Between 18-25 | 40,190 | 16,161 | 40.2% (39.7%-40.7%) | 8.69 (8.46-8.92) |
| Between 25-40 | 145,003 | 63,835 | 44.0% (43.8%-44.3%) | 9.69 (9.49-9.89) |
| Between 40-65 | 140,255 | 85,037 | 60.6% (60.4%-60.9%) | 17.11 (16.75-17.48) |
| Greater than 65 | 35,477 | 26,036 | 73.4% (72.9%-73.8%) | 24.24 (23.49-25.02) |
| **COVID-19 Comorbidities** |  |  |  |  |
| No | 422,116 | 125,996 | 29.8% (29.7%-30.0%) | Reference |
| Yes | 128,215 | 78,620 | 61.3% (61.1%-61.6%) | 1.55 (1.52-1.57) |
| **Province** |  |  |  |  |
| Eastern Cape | 40,877 | 15,837 | 38.7% (38.3%-39.2%) | Reference |
| Free State | 22,347 | 8,363 | 37.4% (36.8%-38.1%) | 1.04 (1.00-1.08) |
| Gauteng | 249,646 | 96,709 | 38.7% (38.5%-38.9%) | 1.18 (1.15-1.21) |
| KwaZulu-Natal | 85,572 | 27,698 | 32.4% (32.1%-32.7%) | 0.79 (0.77-0.81) |
| Limpopo | 19,410 | 5,981 | 30.8% (30.2%-31.5%) | 0.91 (0.87-0.94) |
| Mpumalanga | 18,514 | 5,084 | 27.5% (26.8%-28.1%) | 0.69 (0.66-0.72) |
| North-West | 19,064 | 6,129 | 32.1% (31.5%-32.8%) | 0.86 (0.82-0.89) |
| Northern Cape | 10,002 | 3,358 | 33.6% (32.6%-34.5%) | 0.84 (0.80-0.88) |
| Western Cape | 84,895 | 35,457 | 41.8% (41.4%-42.1%) | 1.08 (1.06-1.12) |
| **Total** | **550,327** | **204,616** | **37.2% (37.1%-37.3%)** |  |

Across the predictor variables, females (39.45%, 95%CI 39.2%, 39.6%), individuals aged 65 and above (73.4%, 95%CI 72.9%, 73.8%), those with 1 or more comorbidity (61.3%, 95%CI 61.1%, 61.6%) and those from the Western Cape (41.8%, 95%CI 41.4%, 42.1%) had higher vaccination rates. In multivariate analysis, after adjustment for all other factors, males were less likely to have vaccinated than females (AOR 0.89, 95%CI 0.88, 0.90), while individuals aged 65 and above (AOR 24.24, 95%CI 23.49, 25.02), those with COVID-19 comorbidities (AOR 1.55, 95%CI 1.52, 1.57) and those from Gauteng (AOR 1.18, 95%CI 1.15, 1.21) were more likely to be vaccinated [Table 2].

Table 3 presents the distribution of the study population by predicted probability of vaccination and compares the observed proportion vaccinated with the model-predicted proportion in each stratum. The predicted probability of vaccination was estimated using logistic regression. Individuals were then grouped into strata based on their predicted probability (e.g., 0–10%, 10–20%, etc.), allowing comparison of actual and predicted vaccination rates across groups with similar predicted likelihoods. The table shows, for each predicted probability stratum, the number of individuals in that stratum, the proportion of the total population they represent, the observed vaccination rate, and the model-predicted vaccination rate.

**Table 3.** Observed and predicted vaccination rates by predicted probability strata.

| Predicted strata *** | N | % of total | Observed * | Predicted ** |
| --- | --- | --- | --- | --- |
| 0%-10% | 182,570 | 33.2% | 6.8% | 7.0% |
| 10%-20% | 6,832 | 1.2% | 17.2% | 11.3% |
| 20%-30% | 697 | 0.1% | 23.4% | 29.1% |
| 30%-40% | 49,000 | 8.9% | 35.7% | 36.0% |
| 40%-50% | 126,233 | 22.9% | 44.7% | 44.6% |
| 50%-60% | 77,360 | 14.1% | 56.6% | 56.0% |
| 60%-70% | 67,375 | 12.2% | 65.3% | 64.9% |
| 70%-80% | 40,260 | 7.3% | 72.4% | 74.1% |
| **Grand Total** | **550,327** | **100.0%** | **37.2%** | **37.2%** |

\*Observed: Actual percentage of individuals in each stratum who received at least one COVID-19 vaccine dose during the study period.
\*\*Predicted: Average predicted probability of vaccination for individuals in each stratum, as estimated by the logistic regression model.
\*\*\*Predicted Strata: Individuals are grouped into strata based on their predicted probability of vaccination (estimated from logistic regression using sex, age, comorbidity status, and province). The stratum "0–10%" contains individuals with a predicted probability of 0% to 10%; "10–20%" contains those with predicted probability of 10% to 20%; and so on.

One-third of members (33.2%) fell into the lowest vaccination probability band (0–10%), within which 6.8% were vaccinated, closely aligning with the model’s predicted 7.0%. At the opposite end of the spectrum, among those with a predicted probability of 70–80% (7.3% of the sample), 72.4% were vaccinated, again consistent with the predicted 74.1%. Overall, at the population level, the model performed well, with the observed vaccination proportion of 37.2% exactly matching the predicted 37.2%. This alignment across strata provides confidence in the predictive validity of the model and suggests that the logistic regression adequately captured the main demographic and clinical determinants of vaccination uptake in this insured population. The two smallest strata (10–20% and 20–30%), comprising approximately 1.3% of the sample, showed less precise alignment, likely reflecting small cell sizes. Given the absence of financial or physical access barriers to vaccination during the study period, the close alignment between predicted and observed uptake suggests that persistent non-vaccination among higher-probability groups was primarily due to personal choice rather than constrained circumstance or access barriers.

### Part 2: Excess Costs and Cross-subsidisation

Table 4 compares the observed annual per capita COVID-19–related expenditure for vaccinated and non-vaccinated members with the predicted expenditure if all members had been vaccinated under the universal vaccination scenario, stratified by sex, age, comorbidity status, and province. For all groups, the “Observed” columns show the actual costs recorded in the claims data for members in that category. The “Predicted” columns have different interpretations depending on the group. For non-vaccinated members, the “Predicted” column shows the modelled cost if these individuals had been vaccinated (counterfactual). For vaccinated members, the “Predicted” column shows model-predicted costs based on the vaccinated cohort, serving as a validation check. The “All members” columns present the weighted average expenditure across the entire population, with the predicted column showing the counterfactual scenario where all members are vaccinated.

**Table 4.**
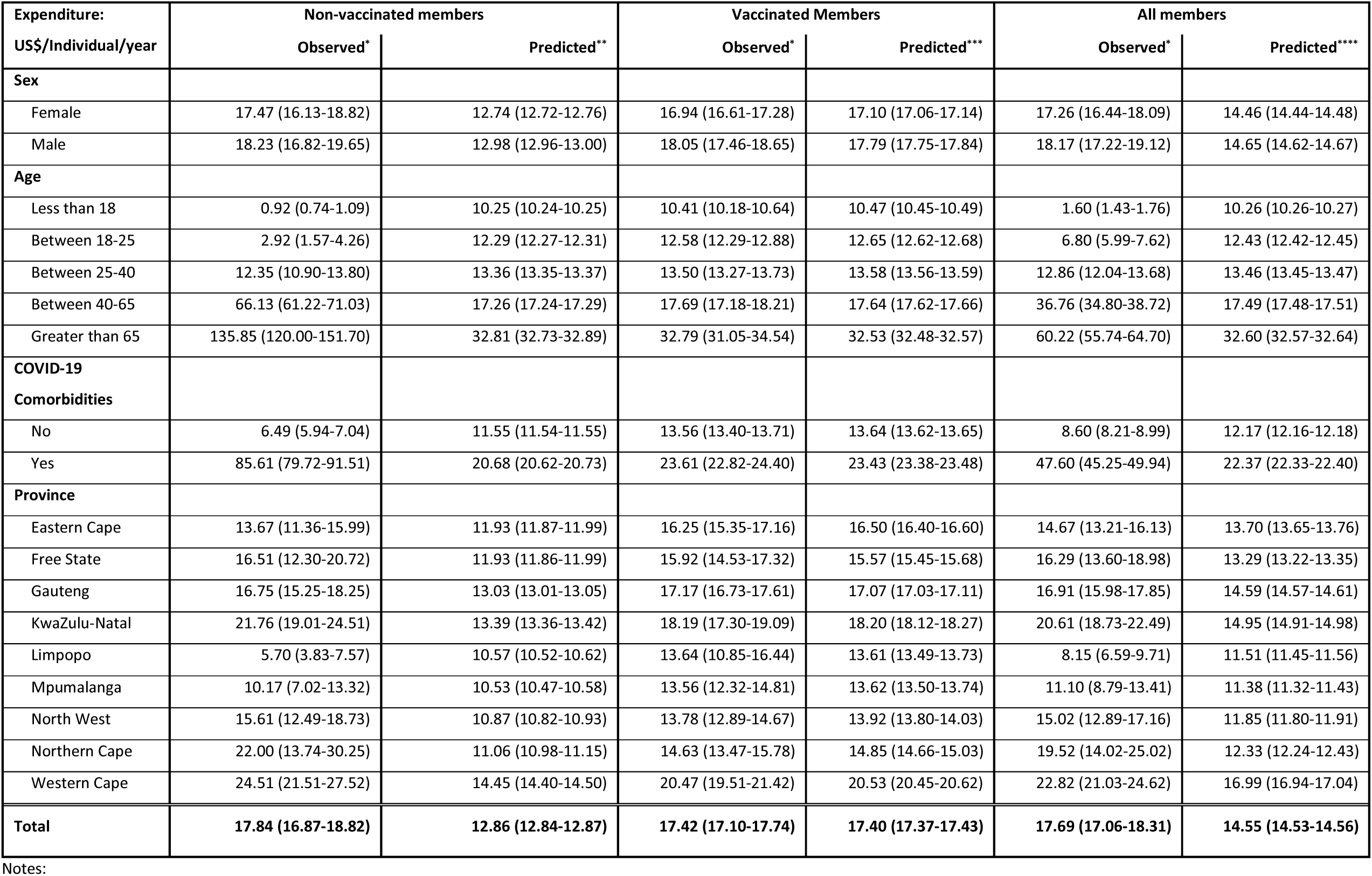
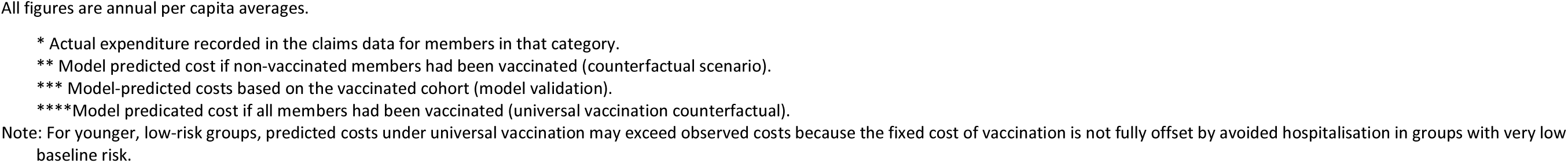
Observed and predicted annual per capita COVID-19 expenditure (US$) by vaccination status and population subgroup.

| Expenditure:<br>US\$/Individual/year | Non-vaccinated members | | Vaccinated Members | | All members | |
| --- | --- | --- | --- | --- | --- | --- |
|  | Observed* | Predicted** | Observed* | Predicted*** | Observed* | Predicted**** |
| **Sex** |  |  |  |  |  |  |
| Female | 17.47 (16.13-18.82) | 12.74 (12.72-12.76) | 16.94 (16.61-17.28) | 17.10 (17.06-17.14) | 17.26 (16.44-18.09) | 14.46 (14.44-14.48) |
| Male | 18.23 (16.82-19.65) | 12.98 (12.96-13.00) | 18.05 (17.46-18.65) | 17.79 (17.75-17.84) | 18.17 (17.22-19.12) | 14.65 (14.62-14.67) |
| **Age** |  |  |  |  |  |  |
| Less than 18 | 0.92 (0.74-1.09) | 10.25 (10.24-10.25) | 10.41 (10.18-10.64) | 10.47 (10.45-10.49) | 1.60 (1.43-1.76) | 10.26 (10.26-10.27) |
| Between 18-25 | 2.92 (1.57-4.26) | 12.29 (12.27-12.31) | 12.58 (12.29-12.88) | 12.65 (12.62-12.68) | 6.80 (5.99-7.62) | 12.43 (12.42-12.45) |
| Between 25-40 | 12.35 (10.90-13.80) | 13.36 (13.35-13.37) | 13.50 (13.27-13.73) | 13.58 (13.56-13.59) | 12.86 (12.04-13.68) | 13.46 (13.45-13.47) |
| Between 40-65 | 66.13 (61.22-71.03) | 17.26 (17.24-17.29) | 17.69 (17.18-18.21) | 17.64 (17.62-17.66) | 36.76 (34.80-38.72) | 17.49 (17.48-17.51) |
| Greater than 65 | 135.85 (120.00-151.70) | 32.81 (32.73-32.89) | 32.79 (31.05-34.54) | 32.53 (32.48-32.57) | 60.22 (55.74-64.70) | 32.60 (32.57-32.64) |
| **COVID-19 Comorbidities** |  |  |  |  |  |  |
| No | 6.49 (5.94-7.04) | 11.55 (11.54-11.55) | 13.56 (13.40-13.71) | 13.64 (13.62-13.65) | 8.60 (8.21-8.99) | 12.17 (12.16-12.18) |
| Yes | 85.61 (79.72-91.51) | 20.68 (20.62-20.73) | 23.61 (22.82-24.40) | 23.43 (23.38-23.48) | 47.60 (45.25-49.94) | 22.37 (22.33-22.40) |
| **Province** |  |  |  |  |  |  |
| Eastern Cape | 13.67 (11.36-15.99) | 11.93 (11.87-11.99) | 16.25 (15.35-17.16) | 16.50 (16.40-16.60) | 14.67 (13.21-16.13) | 13.70 (13.65-13.76) |
| Free State | 16.51 (12.30-20.72) | 11.93 (11.86-11.99) | 15.92 (14.53-17.32) | 15.57 (15.45-15.68) | 16.29 (13.60-18.98) | 13.29 (13.22-13.35) |
| Gauteng | 16.75 (15.25-18.25) | 13.03 (13.01-13.05) | 17.17 (16.73-17.61) | 17.07 (17.03-17.11) | 16.91 (15.98-17.85) | 14.59 (14.57-14.61) |
| KwaZulu-Natal | 21.76 (19.01-24.51) | 13.39 (13.36-13.42) | 18.19 (17.30-19.09) | 18.20 (18.12-18.27) | 20.61 (18.73-22.49) | 14.95 (14.91-14.98) |
| Limpopo | 5.70 (3.83-7.57) | 10.57 (10.52-10.62) | 13.64 (10.85-16.44) | 13.61 (13.49-13.73) | 8.15 (6.59-9.71) | 11.51 (11.45-11.56) |
| Mpumalanga | 10.17 (7.02-13.32) | 10.53 (10.47-10.58) | 13.56 (12.32-14.81) | 13.62 (13.50-13.74) | 11.10 (8.79-13.41) | 11.38 (11.32-11.43) |
| North West | 15.61 (12.49-18.73) | 10.87 (10.82-10.93) | 13.78 (12.89-14.67) | 13.92 (13.80-14.03) | 15.02 (12.89-17.16) | 11.85 (11.80-11.91) |
| Northern Cape | 22.00 (13.74-30.25) | 11.06 (10.98-11.15) | 14.63 (13.47-15.78) | 14.85 (14.66-15.03) | 19.52 (14.02-25.02) | 12.33 (12.24-12.43) |
| Western Cape | 24.51 (21.51-27.52) | 14.45 (14.40-14.50) | 20.47 (19.51-21.42) | 20.53 (20.45-20.62) | 22.82 (21.03-24.62) | 16.99 (16.94-17.04) |
| **Total** | **17.84 (16.87-18.82)** | **12.86 (12.84-12.87)** | **17.42 (17.10-17.74)** | **17.40 (17.37-17.43)** | **17.69 (17.06-18.31)** | **14.55 (14.53-14.56)** |

Notes:
\* Actual expenditure recorded in the claims data for members in that category.
\*\* Model predicted cost if non-vaccinated members had been vaccinated (counterfactual scenario).
\*\*\* Model-predicted costs based on the vaccinated cohort (model validation).
\*\*\*\*Model predicated cost if all members had been vaccinated (universal vaccination counterfactual).
Note: For younger, low-risk groups, predicted costs under universal vaccination may exceed observed costs because the fixed cost of vaccination is not fully offset by avoided hospitalisation in groups with very low baseline risk.

The results show that observed and predicted expenditures were closely aligned among vaccinated members (US$17.42 vs. US$17.40), confirming good model calibration. Among non-vaccinated members, observed expenditure exceeded predicted costs overall (US$17.84 vs. US$12.86), particularly in older adults (≥65 years: US$135.85 vs. US$32.81) and those with comorbidities (US$85.61 vs. US$20.68), indicating substantial excess costs attributable to non-vaccination in higher-risk groups. In contrast, among younger members (<40 years), predicted costs under a universal vaccination scenario were slightly higher than observed costs, reflecting that vaccination added a modest direct cost in groups with very low hospitalisation risk. These dynamics are further considered in the discussion section.

Table 5 presents the comparison between observed and predicted annual COVID-19–related expenditure per individual, stratified by predicted probability of vaccination. All figures are annual per capita averages. The excess cost column shows the difference between observed and predicted expenditure (observed minus predicted) for all members in each stratum. Although the calculation includes all members, the excess cost essentially reflects the difference attributable to non-vaccinated members, as observed and predicted costs for vaccinated members were very close (US$17.43 vs. US$17.41 overall). Positive values indicate that observed costs exceeded predicted costs under universal vaccination, as seen among non-vaccinated members in higher-probability strata. Negative values indicate that predicted costs exceeded observed spending.

**Table 5.**
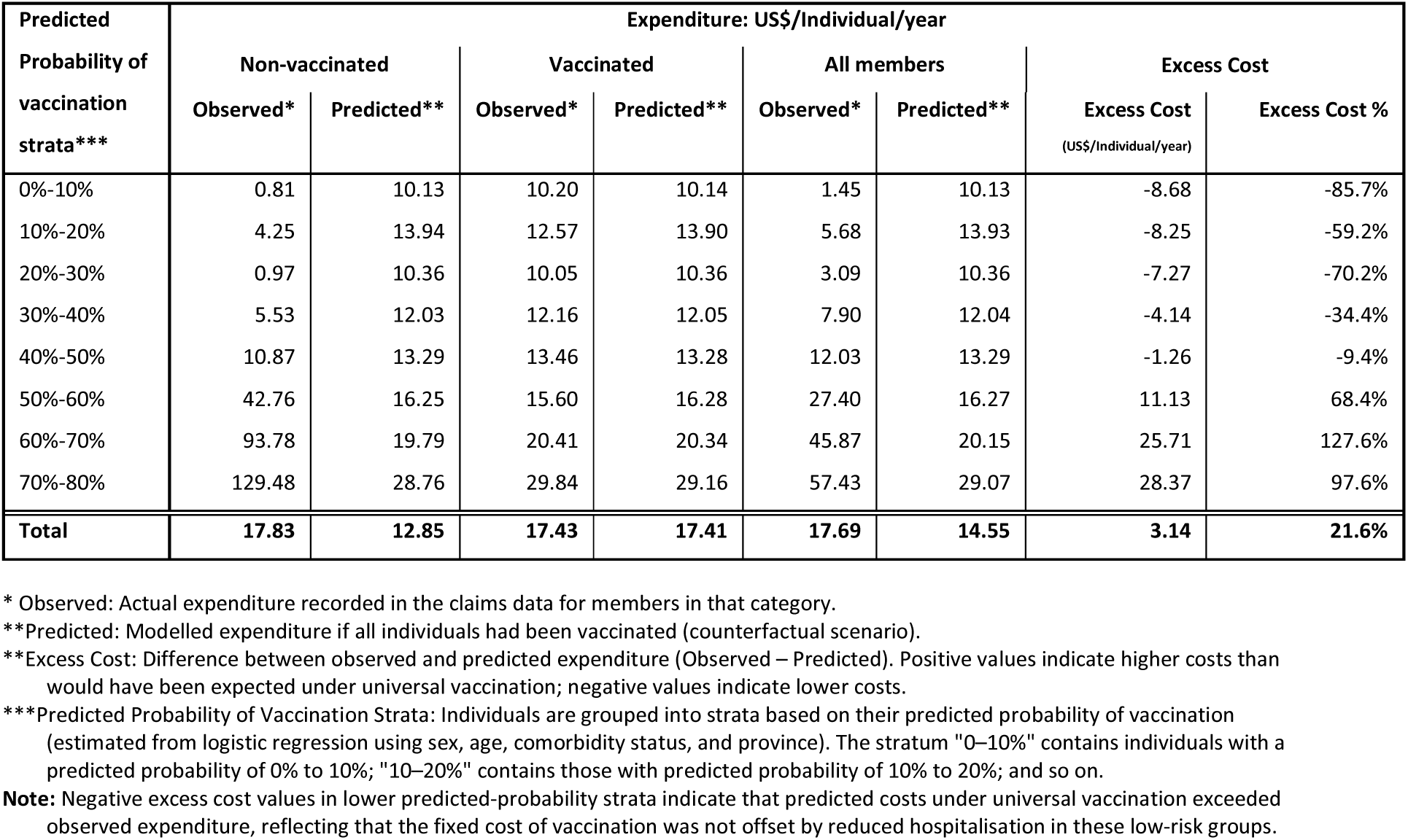
Observed and predicted expenditure (US$) by predicted probability of vaccination strata.

Among members with low predicted vaccination probability (<30%), observed expenditure was minimal, ranging from US$0.81 to US$4.25 across strata, and substantially below predicted costs under a universal vaccination scenario (US$10.13– US$13.94). This resulted in negative excess cost values (–US$7.27 to –US$8.68) and corresponding excess cost percentages of –70.2% to –85.7%. This indicates that in low-predicted probability of vaccination strata; comprising largely younger, low-risk members; the direct costs of vaccination would have exceeded the limited costs associated with hospitalisation.

From the 30–40% predicted vaccination probability band upward, however, the relationship reversed. Observed expenditure among non-vaccinated members rose steeply relative to predicted costs, with excess costs increasing progressively across higher predicted vaccination probability strata. In the 50–60% band, non-vaccinated members incurred mean costs of US$42.76 versus a predicted US$16.30 (excess cost = US$11.13; +68.4%). The disparity widened further in the 60–70% and 70–80% bands, where observed costs were four to five times higher than predicted (US$93.78 vs. US$19.8 and US$129.48 vs. US$28.76, respectively), corresponding to excess burdens of +127.6% and +97.6%.

At the population level, non-vaccinated members had observed average costs of US$17.83 compared to a predicted average of US$12.85, yielding an overall excess burden of US$3.14 per individual per year (+21.6%). For vaccinated members, observed and predicted expenditures were nearly identical (US$17.43 vs. US$17.41), confirming the validity of the cost model. Collectively, these results show that excess costs from non-vaccination were concentrated among individuals with a high predicted likelihood of vaccination, those for whom non-vaccination was more likely a matter of personal choice rather than constrained circumstance.

## DISCUSSION

This study applied a luck egalitarian framework to examine COVID-19 vaccination uptake and associated costs within a large South African insured population. By combining a contextual review of the policy environment and communications and regulatory requirements with empirical predictive modelling, the study sought to distinguish non-vaccination due to personal choice from non-uptake due to constrained circumstance, including potential access barriers, and to quantify the financial implications of these behavioural differences for the insurance risk pool. In doing so, it responds to the broader debate about whether, and under what conditions, personal responsibility should inform health financing decisions in systems pursuing Universal Health Coverage.

Two main findings emerge from the study. First, non-vaccination in this insured population was unlikely to result from financial or physical access barriers, given the absence of user fees for vaccination, widespread availability of vaccination sites, and the extent of public and insurer-level communication. Predictive modelling, validated by the close alignment between predicted and actual uptake, provided a basis for inferring that non-vaccination in higher-probability groups plausibly reflected personal choice.

Second, unvaccinated members generated measurable excess costs relative to modelled expectations. When expenditure patterns from vaccinated members were applied across the entire population, observed costs exceeded predicted costs by an average of 22%. These excess costs were concentrated among individuals with high predicted vaccination probabilities, those who had the highest risk factors for severe COVID-19 and for whom non-vaccination is most plausibly due to personal choice. In the 50–60% probability band, observed costs exceeded predicted costs by 68.4%. The disparity widened further in the 60–70% and 70–80% bands, where observed costs exceeded predicted costs by 127.6% and 97.6%, respectively.

Among younger, low-risk members in the lowest probability strata (<30%), however, predicted costs under universal vaccination exceeded observed expenditure. For example, in the 0–10% band, observed costs were 92% lower than predicted costs. This reflects that the fixed cost of vaccination was not fully offset by avoided hospitalisation in groups with very low baseline risk. This reversal illustrates the efficiency–equity trade-off inherent in preventive interventions: vaccination yields modest direct financial benefits for low-risk individuals but substantially reduces costs and severe outcomes among higher-risk groups, strengthening the equity and sustainability of the collective risk pool. Taken together, these findings highlight the tensions between responsibility-sensitive fairness and financial solidarity in private health insurance arrangements.

Much of the existing literature on luck egalitarianism in health care is normative and conceptual. This study provides an empirical demonstration of how these principles can be operationalised in a real-world context. The central luck egalitarian claim that inequalities traceable to “brute luck” should be compensated, while those resulting from option luck may legitimately be left for individuals to bear has been influential in health ethics (Dworkin, 1981a, Arneson, 1989, Cohen, 2011). Scholars such as Segall (Segall, 2010)and Knight (Knight, 2009) have examined the implications of responsibility-sensitive allocation in health systems, while Albertsen and Knight(Albertsen et al., 2014) explored practical limits.

Our analysis contributes by showing that in this insured South African cohort, it was possible to contextually establish that access barriers were minimal, to identify individuals for whom non-vaccination could reasonably be attributed to personal choice, and to quantify the excess costs imposed on the risk pool by these individuals. These findings align with arguments that personal responsibility may be a legitimate distributive principle when genuine personal choice exists (Voigt, Buyx, 2008). At the same time, we recognize the critiques of responsibility-based approaches. Social determinants, misinformation, and systemic distrust may have influenced vaccine hesitancy in ways not fully observable in claims data (Wikler, 2002, Brown et al., 2019). Other empirical and theoretical studies have highlighted how such unmeasured contextual factors complicate straightforward attribution of responsibility (Albertsen, 2019, Levy et al., 2011) and risk reinforcing disadvantage if overlooked.

The observed cross-subsidisation has direct implications for health financing and fairness within insured risk pools. In a context where premiums are community-rated, vaccinated members effectively cross-subsidised the higher costs incurred by unvaccinated members, raising questions about whether differentiated cost-sharing or incentives could be justified, consistent with luck egalitarian reasoning (Cutler et al., 2000, Segall, 2010). However, implementing responsibility-sensitive policies requires caution to avoid unfairly penalising individuals whose non-vaccination may still reflect unmeasured barriers. More broadly, the findings highlight the tension between solidarity, efficiency, and responsibility in health financing arrangements (Albertsen et al., 2014). While we do not argue for punitive policies against the unvaccinated, our results highlight the need for transparent debate about the fairness of cross-subsidies when risks are voluntarily assumed, and the conditions under which responsibility may legitimately be invoked.

This study has several limitations. First, the findings are specific to COVID-19 vaccination in an insured South African population and cannot be generalised to uninsured populations or different health system contexts. Second, the analysis uses routine “real world” demographic and claims data collected by the health insurance funds as part of their business, not data collected for research purposes per se. Only services and vaccines for which claims were submitted were analysed. The implications are that both COVID-19 vaccination events and hospitalization for COVID-19 infection could be over or underreported. While it is difficult to predict the impact of these biases, it seems plausible that they would not negate the findings entirely, given the magnitude of the impact. Third, the counterfactual cost model assumes that the relationship between covariates and costs is the same for vaccinated and unvaccinated individuals. If unvaccinated members differed systematically in unobserved ways, for instance, in health-seeking behaviour or adherence to other preventive measures, the predicted costs under universal vaccination may be biased. Fourth, the binary end-of-period vaccination classification does not capture variation in the timing of vaccination relative to COVID-19 waves, which may affect both the choice inference and the cost estimates. Fifth, we lacked individual-level data on geographic availability (distance to vaccination sites, waiting times) and acceptability (vaccine hesitancy, trust, misinformation). These unmeasured factors may have influenced vaccination decisions in ways not captured by our analysis, and caution is warranted in interpreting residual non-vaccination as solely a matter of personal choice.

## CONCLUSION

This study set out to examine whether non-vaccination for COVID-19 in a South African insured population reflected personal choice or constrained circumstance, and to quantify the resulting financial implications for the insurance risk pool. Within a context of comprehensive vaccine availability, minimal access barriers, and extensive communication, non-vaccination was more plausibly attributable to personal choice than to constrained circumstance.

Using predictive and expenditure modelling, we found that observed costs exceeded predicted costs by an average of 22% under a universal vaccination scenario, with the excess concentrated among those whose non-vaccination most plausibly reflected personal choice. Among younger, low-risk members, the fixed cost of vaccination was not fully offset by avoided hospitalisation. The study demonstrates that non-vaccination due to personal choice can be empirically distinguished from non-vaccination due to constrained circumstance, and that the financial consequences of that distinction can be quantified. By operationalising luck egalitarian principles in a real-world context, this research offers an empirical basis for deliberating when and how personal responsibility should inform the design of equitable and sustainable health systems.

## Declaration of generative AI and AI and AI assisted technologies in the manuscript preparation process

During the preparation of this work the author(s) used ChatGPT and DeepSeek order to to assist with language editing, phrasing refinement, and structural clarity. After using this tool/service, the author(s) reviewed and edited the content as needed and take(s) full responsibility for the content of the published article.

## Ethical approval

Ethics approval was granted by the Faculty of Health Sciences Human Research Ethics Committee of the University of Cape Town – approval number HREC Ref: 606/2023.

## Author contributions

Geetesh Solanki, Susan Cleary and Francesca Little designed the study. Geetesh Solanki prepared and ran the analytical models with input from Francesca Little. Susan Cleary provided critical review oversight. Geetesh Solanki wrote the first draft of the manuscript which was then critically reviewed and revised by the other authors.

## Data Availability

De-identified data supporting the findings of this study are available on ZivaHub, the University of Cape Town’s institutional research data repository, at https://doi.org/10.25375/uct.30533219. The dataset includes anonymised demographic, vaccination, and claims expenditure records used in the analyses. Data are shared under a Creative Commons Attribution (CC BY 4.0) license and may be reused for non-commercial research with appropriate citation.

## Acknowledgements

The authors would like to acknowledge the institutional support to the study authors provided by the University of Cape Town. We also acknowledge NMG Consultant and Actuaries for assisting in providing access to the data and Priya Makanjee for assistance in extracting the data.

